# Epidemiology and excess mortality of antimicrobial resistance in bacteraemias among cancer patients: a cohort study using routinely collected health data from regional hospital trusts in Oxford and Oslo, 2008-2018

**DOI:** 10.1101/2024.08.21.24312363

**Authors:** Anders Skyrud Danielsen, Cherry Lim, Chang Ho Yoon, Jon Michael Gran, Oliver Kacelnik, David W. Eyre, Jørgen Vildershøj Bjørnholt

## Abstract

**Objectives:** We investigated the epidemiology and impact on mortality of antimicrobial resistance (AMR) in cancer patients with bacteraemia at Oxford University Hospitals (OxUH), UK, and Oslo University Hospital (OsUH), Norway, during 2008-2018.

**Design:** Historical cohort study.

**Setting:** Regional hospital trusts with multiple sites in OxUH and OsUH.

**Methods:** Patients with cancer and blood cultures positive for one of six pathogen groups during a hospital stay within three years following their first cancer diagnosis were followed for 30 days after their first bacteraemia episode. We determined the number of cases and the proportion of infections with an AMR phenotype. Excess mortality and the population-attributable fraction (PAF) due to AMR were estimated by contrasting observed mortality at the end of follow-up with an estimated counterfactual scenario where AMR was absent from all bacteraemias, using inverse probability weighting.

Main outcome measure: 30-day all-cause mortality following the first bacteraemia episode. Main exposure measure: A resistant phenotype of the causative pathogen.

**Results:** The study included 1929 patients at OxUH and 1640 patients at OsUH. The highest resistance proportions were found for vancomycin-resistance in enterococci (85/314, 27.1%) and carbapenem-resistance in *Pseudomonas aeruginosa* (63/260, 24.2%) at OxUH, and third-generation cephalosporin-resistance in *Escherichia coli* (62/743, 8.3%) and *Klebsiella pneumoniae* (14/223, 6.3%) at OsUH. Observed mortality for all infections was 26.4% at OxUH, with an estimated counterfactual mortality without AMR of 24.7%, yielding an excess mortality of 1.7% (95% CI: 0.8-2.5%). The PAF was 6.3% (95% CI: 2.9-9.6%), meaning an estimated 32 of 509 deaths could be attributed to AMR. Limited events at OsUH precluded a similar estimate.

**Conclusions:** Despite estimating modest excess mortality, the mortality attributable to resistance in these two high-income, low-prevalence settings highlights the potential for escalation if global resistance trends continue to worsen.

## Introduction

As antimicrobial resistance (AMR) is growing in prevalence, precise estimation and surveillance of its disease burden is critical [1]. The excess mortality that can be attributed to AMR has been estimated using a wide range of methodologies [1–5]. Such estimations may inform healthcare policies and guide clinical decision-making to prevent deaths. Despite the recognised need, estimating excess mortality from observational data remains methodologically challenging and has produced conflicting results [6–8]. However, a counterfactual approach is generally considered necessary for estimating attributable mortality.

Cancer patients are particularly vulnerable to infections due to the immunosuppressive nature of both disease and treatment [9–11], and the presence of AMR in infectious organisms can significantly exacerbate the risks of adverse outcomes [9]. Especially, the safeguarding of high-risk cancer treatments such as stem cell transplantation, radical resections, or cytotoxic drugs requires an understanding of the implications of AMR within these patient populations.

In this study, we aimed to describe the microbial epidemiology of bacteraemias in cancer patients at two large regional hospital trusts with multiple sites – namely Oxford University Hospitals (OxUH) in the United Kingdom and Oslo University Hospital (OsUH) in Norway – from 2008 to 2018. We focused on eight key drug-pathogen combinations that have been shown to be important in Europe [2,3]. Additionally, we aimed to estimate the excess mortality attributable to AMR at the end of follow-up utilising a counterfactual framework.

## Methods

### Study design and cohort selection

We used routinely collected health data to create two cohorts of patients with cancer who had a positive blood culture (bacteraemia) during a hospital stay within three years following their first cancer diagnosis at OxUH and OsUH between 2008 and 2018 (Table 1, Figure 1). Patients were included at the sample date and time of their first positive blood culture during the study period (day 0) and followed for 30 days for all-cause mortality. Only the first bacteraemia episode per patient was considered, ensuring each patient was represented only once.

**Table 1.** Study design elements.

| Study design element | Description |
| --- | --- |
| Eligibility criteria | Patients with cancer who had a positive blood culture (bacteraemia) within three years following their first cancer diagnosis at Oxford University Hospitals or Oslo University Hospital between 2008 and 2018. |
| Follow-up period | The start of follow-up was the date and time of the first positive blood culture within the study period and patients were followed for 30 days. |
| Exposure groups | Group 1: Patients with bacteraemia caused by bacteria susceptible to antibiotics from the selected drug-pathogen combinations.<br>Group 2: Patients with bacteraemia caused by bacteria resistant to antibiotics from the selected drug-pathogen combinations. |
| Estimand | The difference between:<br>- the observed 30-day all-cause mortality for both groups and<br>- the counterfactual (hypothetical) 30-day all-cause mortality if all bacteraemia cases had been caused by susceptible bacteria. |

**Figure 1.**
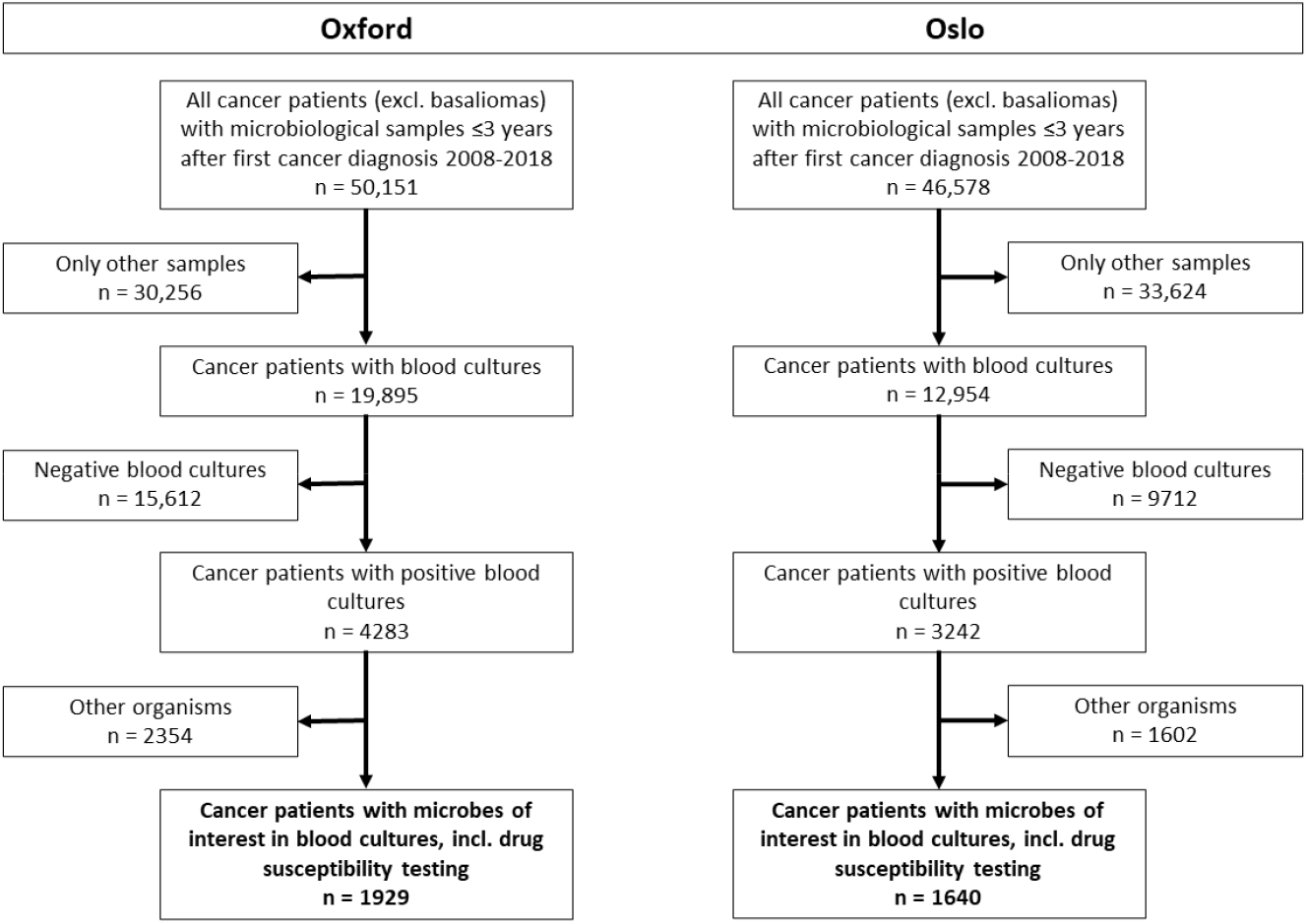
Flow chart of the cohort selection process. “Other organisms” were organisms other than those included in the eight drug-pathogen combinations.

Patients were divided into two groups based on the AMR phenotype of the infectious organism isolated in the bacteraemia. We focused on six pathogen groups: *Staphylococcus aureus, Enterococcus faecalis* and *Enterococcus faecium* (enterococci), *Escherichia coli, Klebsiella pneumoniae, Pseudomonas aeruginosa*, and *Acinetobacter* spp. These pathogens were then subclassified as having either a resistant or susceptible phenotype according to EUCAST breakpoints, specifically against methicillin (methicillin-resistant *S. aureus*, MRSA), vancomycin (vancomycin-resistant enterococci, VRE), third generation cephalosporins or carbapenems (third generation cephalosporin-resistant *E. coli*, 3GCREC; third generation cephalosporin-resistant *K. pneumoniae*, 3GCRKP; carbapenem-resistant *E. coli*, CREC; carbapenem-resistant *K. pneumoniae*, CRKP), and carbapenems alone (carbapenem-resistant *P. aeruginosa*, CRPA; and carbapenem-resistant Acinetobacter spp., CRA), respectively [12]. These classifications resulted in eight specific drug-pathogen combinations for analysis. The selection of these drug-pathogen combinations was based on their contribution to the overall AMR burden in Europe [2,3]. A cumulative antibiogram was also constructed for each included microorganism by assessing resistance across a range of antimicrobials in all microbiological sources. In polymicrobial bacteraemias, cases were defined as resistant if any of the causative agents had discordant resistance profiles or were resistant to at least one of the relevant antibiotics.

Data from OxUH was obtained from the Infections in Oxfordshire Research Database (IORD) with approvals from the National Research Ethics Service South Central-Oxford C Research Ethics Committee (19/SC/0403), Health Research Authority and Confidentiality Advisory Group (19/CAG/0144) as a deidentified database without individual consent. Norwegian registry data retrieval was approved by the Regional Ethics Committee of South East Norway (240258) and the Data Protection Officer at OsUH (21/06874).

### Data sources

For the OxUH cohort, data were sourced from the Infections in Oxfordshire Research Database (IORD), which includes hospital episode data with diagnostic and procedure codes and in-hospital microbiological laboratory data. For the OsUH cohort, data were obtained from the Cancer Registry of Norway (CRN), the Norwegian Patient Registry (NPR), the Cause of Death Registry (CDR), and in-hospital microbiological laboratory data.

### Exposure

The exposure variable was the AMR phenotype of the causative organism in the bacteraemia, classified according to EUCAST breakpoints.

### Outcome

The primary outcome was 30-day all-cause mortality.

### Covariates

Covariate adjustment was informed by a directed acyclic graph (DAG, Supplementary material S1). Covariates included:

- Sex: Recorded as either female or male.
- Age at cancer diagnosis: Treated as a continuous variable, derived from the date of birth.
- Charlson Comorbidity Index (CCI): Calculated from ICD-10 diagnosis codes within one year prior to the bacteraemia episode, reflecting the burden of comorbid conditions and severity of illness, which may influence the likelihood of receiving broad-spectrum antibiotics.
- Cumulative days of hospitalisation in the past year: Sum of all hospital stay days within one year prior to the bacteraemia episode, indicating prior exposure to secondary healthcare.
- Other infectious focus: Presence of infectious syndromes in sites other than blood, identified through ICD-10 codes.
- Year and month of cancer diagnosis: Used to adjust for secular trends, including changes in cancer treatment, resistance patterns, and supportive (sepsis) care over time.
- Cancer type and treatment: Categorised based on malignancy and treatment within a year before follow-up, including solid cancers with or without surgery and haematological cancers with or without stem cell transplantation.
- Polymicrobial bacteraemia: Defined by the identification of different causative agents in the same or adjacent days’ blood cultures, excluding common contaminants like coagulase-negative staphylococci.

A detailed description of the covariates and data sources can be found in Supplementary material S2.

### Statistical analysis

We estimated excess mortality due to AMR as the difference between the observed mortality and the expected mortality had all bacteraemias been caused by susceptible bacteria. The expected mortality was estimated using inverse probability weighting, conditioned on the adjustment set identified through the DAG [13,14].

Multivariable logistic regression was used to calculate stabilised, untruncated inverse probability weights, with the set of covariates as independent variables and the exposure group as the dependent variable. The expected cumulative incidence of mortality was subtracted from the observed cumulative incidence to obtain the excess mortality estimate. Confidence intervals for excess mortality and the population-attributable fraction (PAF) were calculated using nonparametric bootstrapping with 1000 samples.

Kaplan-Meier survival curves were estimated by exposure groups and for the overall observed mortality and for the (weighted) counterfactual scenario. Excess mortality and PAF were illustrated with density plots and bootstrapped confidence intervals [13]. Sensitivity analyses were performed for each drug-pathogen combination and included an additional linear term for cumulative days on antibiotics in the previous year. The E-value was calculated to assess the minimum strength of association required for an unmeasured confounder to explain the estimated excess mortality due to AMR [15].

A detailed description of the statistical analysis can be found in the Supplementary material S3. All statistical analyses were performed using R (version 4.3.0) and the script is available online [16,17].

## Results

We included 1929 patients at OxUH and 1640 at OsUH, diagnosed with cancer and experiencing bacteraemia within three years of their first cancer diagnosis (Figure 1, Table 2). At OxUH, 1620/1929 (84%) had bacteraemia with a susceptible AMR phenotype, and 309/1929 (16%) had bacteraemia with AMR. At OsUH, 1541/1640 (94%) had bacteraemia without AMR and 99/1640 (6%) had bacteraemia with AMR. The median age at OxUH was 70 years (IQR 58-79) for patients without AMR and 66 years (IQR 55-76) for those with AMR, while at OsUH, patients without AMR had a median age of 66 years (IQR 55-75), with those with AMR being younger at a median of 59 years (IQR 44-68). The most common cancer type and treatment at both OxUH and OsUH was solid tumours treated with surgery, comprising 1070/1929 (55%) and 909/1640 (55%) patients, respectively. At OxUH, the proportion of bacteraemias that were categorised as polymicrobial was 76/1929 (4%), with 248/1640 (15%) at OsUH. The bacteraemia was associated with a known focus or clinical syndrome in 810/1620 (50%) of those without AMR and 131/309 (42%) of those with AMR at OxUH, and 649/1541 (42%) and 41/99 (41%) at OsUH, respectively. Patients without AMR had a cumulative median stay of 18 days (IQR 8-35) in hospital for a year prior to bacteraemia onset and those with AMR had 29 days (IQR 15-53) at OxUH, whereas OsUH patients without AMR had a cumulative median stay of 40 days (IQR 21-68) and those with AMR had 57 days (IQR 30-90).

**Table 2.**
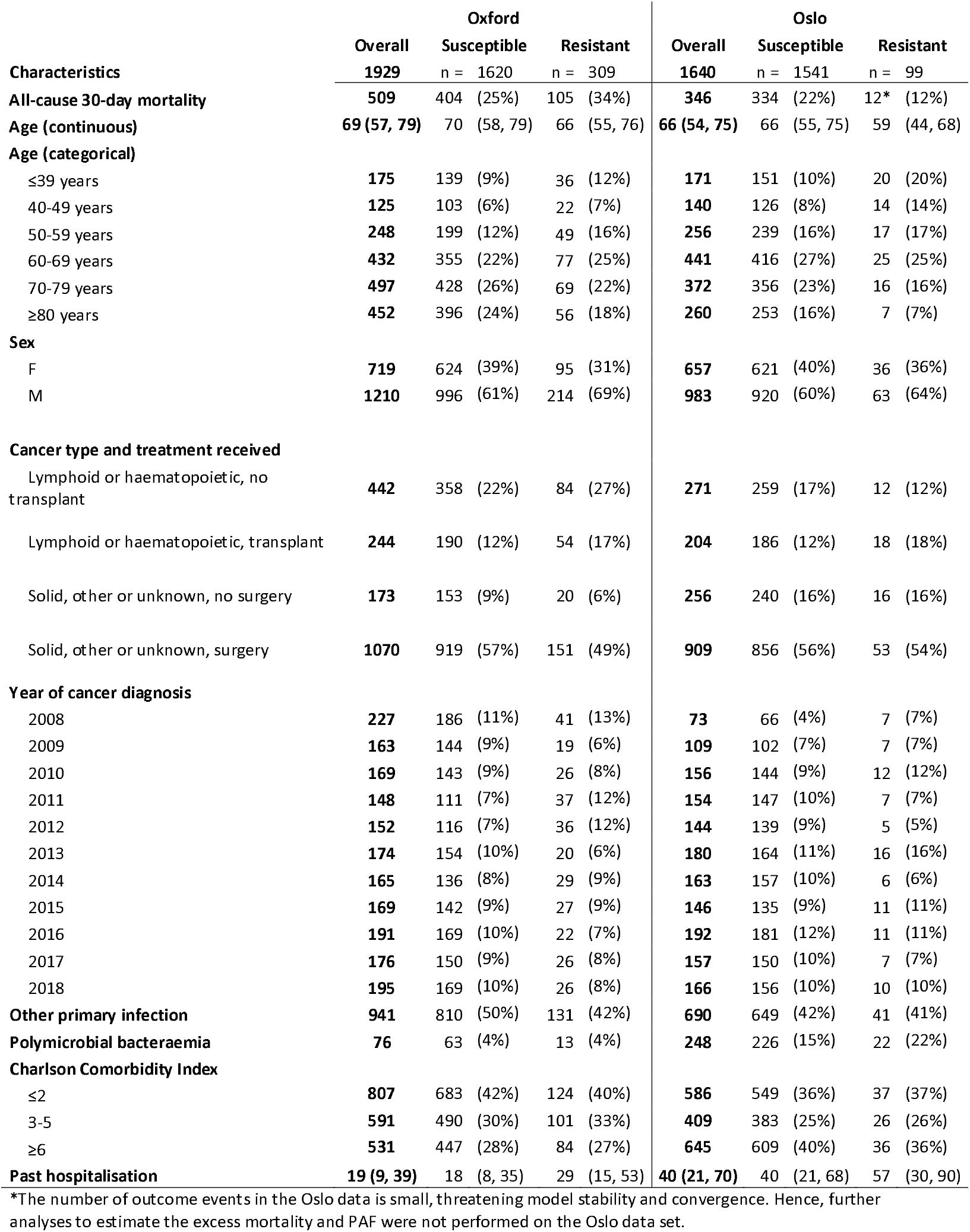
Characteristics of cancer patients diagnosed in 2008-2018 and followed three years after their first cancer diagnosis at Oxford University Hospitals and Oslo University Hospital, categorised by the antimicrobial resistance phenotype of their first bacteraemia episode in the 30-day follow-up period. Continuous covariates are presented with median and interquartile range, while categorical covariates are presented with frequencies and percentages.

Table 3 presents the number of cases and resistance proportions of the drug-pathogen combinations in the bacteraemia episodes at OxUH and OsUH. OxUH had a higher proportion of MRSA among *S. aureus* bacteraemia cases at 13.5% (41/303), and 27.1% (85/314) of enterococci cases were vancomycin resistant (VRE), while OsUH had 3.7% (10/271) and 1.5% (5/342), respectively. Both the cases of P. aeruginosa infection (260) and the proportion resistant to carbapenems (24.2%, 63/260) were higher at OxUH while OsUH had 87 and 3.4% (3/87), respectively. Additionally, the proportion of *Acinetobacter* spp. resistant to carbapenems were 16.7% (6/36) at OxUH, while no resistant strains were reported among the 9 isolates at OsUH.

**Table 3.** Number of cases and resistance proportions of the key drug-pathogen combinations isolated from the blood of cancer patients diagnosed in 2008-2018 at Oxford University Hospitals and Oslo University Hospital.

|  | Oxford |  |  | Oslo |  |  |
| --- | --- | --- | --- | --- | --- | --- |
|  | Resistant | Susceptible | Proportion | Resistant | Susceptible | Proportion |
| *Escherichia coli*, third-generation cephalosporin-resistant | 89 | 870 | 9.3 % | 62 | 681 | 8.3 % |
| *Staphylococcus aureus*, methicillin-resistant | 41 | 262 | 13.5 % | 10 | 261 | 3.7 % |
| *Pseudomonas aeruginosa*, carbapenem-resistant | 63 | 197 | 24.2 % | 3 | 84 | 3.4 % |
| *Klebsiella pneumoniae*, third-generation cephalosporin-resistant | 30 | 205 | 12.8 % | 14 | 209 | 6.3 % |
| *Acinetobacter* spp., carbapenem-resistant | 6 | 30 | 16.7 % | 0 | 9 | 0.0 % |
| *Klebsiella pneumoniae*, carbapenem-resistant | 1 | 231 | 0.4 % | 6 | 216 | 2.7 % |
| Enterococci, vancomycin-resistant | 85 | 229 | 27.1 % | 5 | 337 | 1.5 % |
| *Escherichia coli*, carbapenem-resistant | 5 | 947 | 0.5 % | 4 | 740 | 0.5 % |

The distribution of weights used for inverse probability weighting in the OxUH cohort can be found in Figure 2. The mean of the weights approached 1 as expected: 0.99, a standard deviation of 0.43 and a range of 0.24-2.45 for those with AMR, and a mean of 1.00, a standard deviation of 0.12 and a range of 0.88-2.51 for those without AMR. The unweighted and weighted mean of the continuous and dichotomous covariates included in the weighting model can be found in Supplementary material S3.

**Figure 2.**
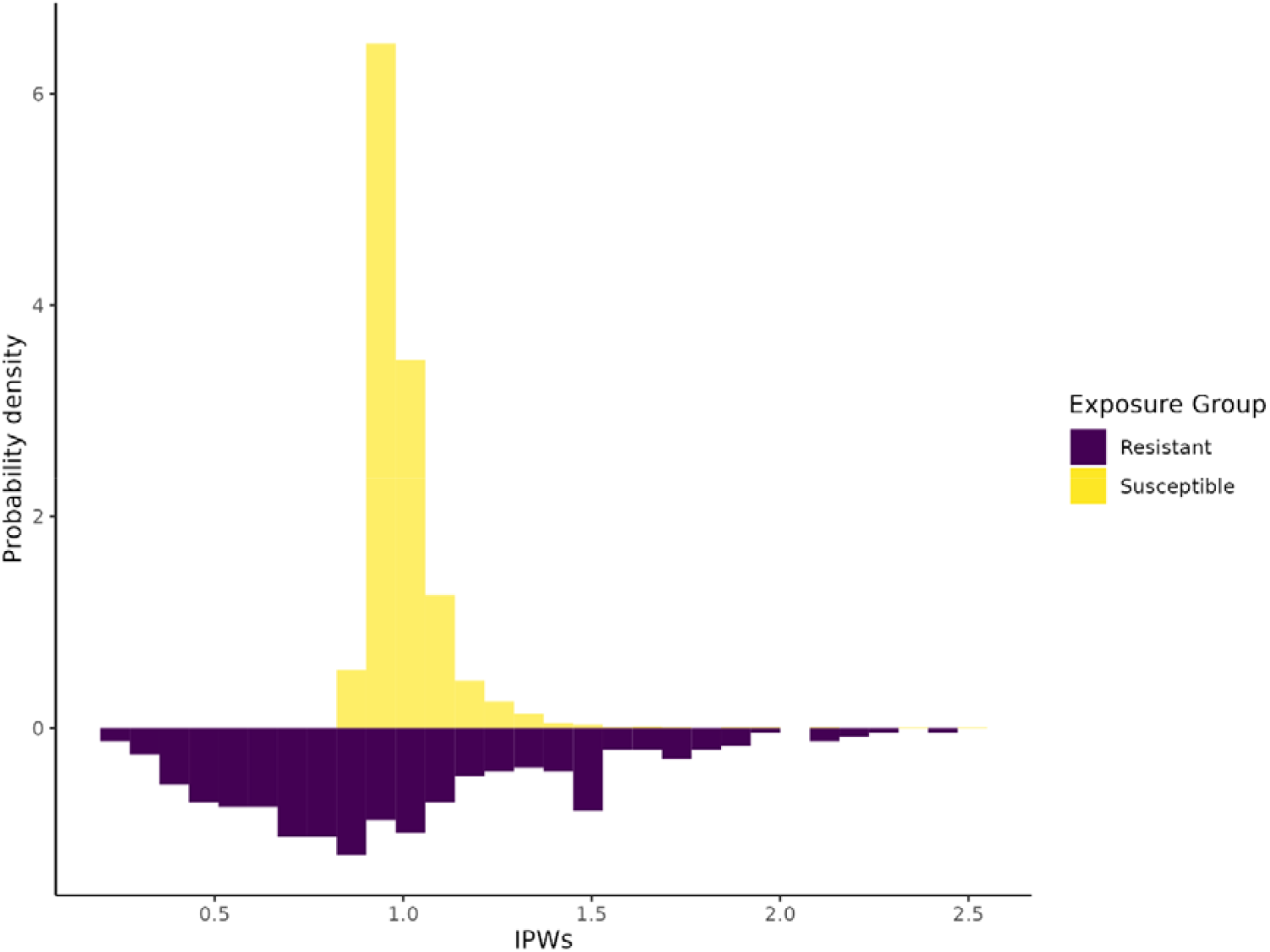
Mirror histograms showing the probability density of the inverse probability weights (IPWs) for each antimicrobial resistance phenotype of the first bacteraemia of cancer patients diagnosed at Oxford University Hospitals 2008-2018 and followed for three years.

Figures 3A and 3B illustrate the crude survival probabilities for cancer patients at OxUH and OsUH, respectively, following their first bacteraemia episode, while Figure 3C displays the survival probabilities of the overall observed population alongside the weighted survival probabilities of the counterfactual scenario, quantified day by day. Figure 3A showed a slight decrease in observed 30-day survival for patients with AMR (105/309, 34%) compared to those without AMR (404/1620, 25%) at OxUH, with a reverse relationship in Figure 3B for patients at OsUH with a lower survival in those without AMR (334/1541, 22%) than those with AMR (12/99, 12%).

**Figure 3.**
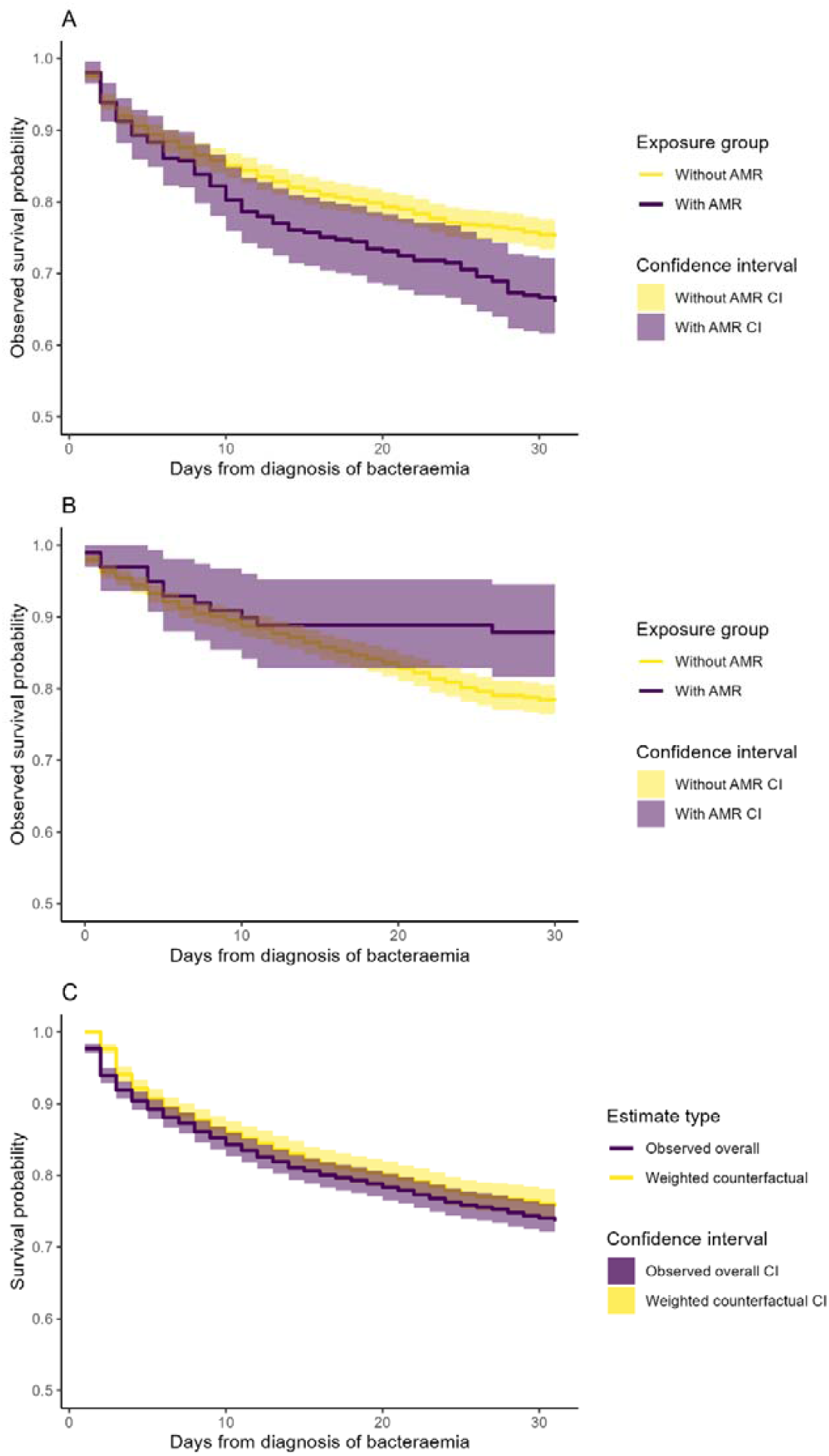
The crude Kaplan-Meier survival curves stratified by the antimicrobial resistance phenotype of the first bacteraemia of cancer patients followed for three years after diagnosis at (A) Oxford University Hospitals and (B) Oslo University Hospital in 2008-2018, and (C) the observed overall survival curve and weighted counterfactual survival curve at Oxford University Hospitals.

The overall observed mortality at the end of follow-up at OxUH was 26.4%, while the estimated mortality under the counterfactual scenario was 24.7%, yielding an excess mortality due to AMR at the end of follow-up of 1.7%, with the bootstrapped 95% CI ranging from 0.8-2.5% (Figure 4). The PAF was estimated at 6.3%, with a bootstrapped 95% CI of 2.9-9.6%, suggesting that approximately 32 of the 509 observed deaths were attributable to AMR. However, due to too few events, it was not feasible to perform a similar estimation for the OsUH cohort. The sensitivity analysis where cumulative previous antibiotic use was added as a linear term gave an unchanged excess mortality of 1.7% (95% CI: 0.8-2.5%. A table with head-to-head comparisons for each of the eight drug-pathogen combinations can be found in Supplementary material S4. A detailed cumulative antibiogram describing the distribution of resistance per pathogen and antimicrobial agent is provided in Supplementary material S5.

**Figure 4.**
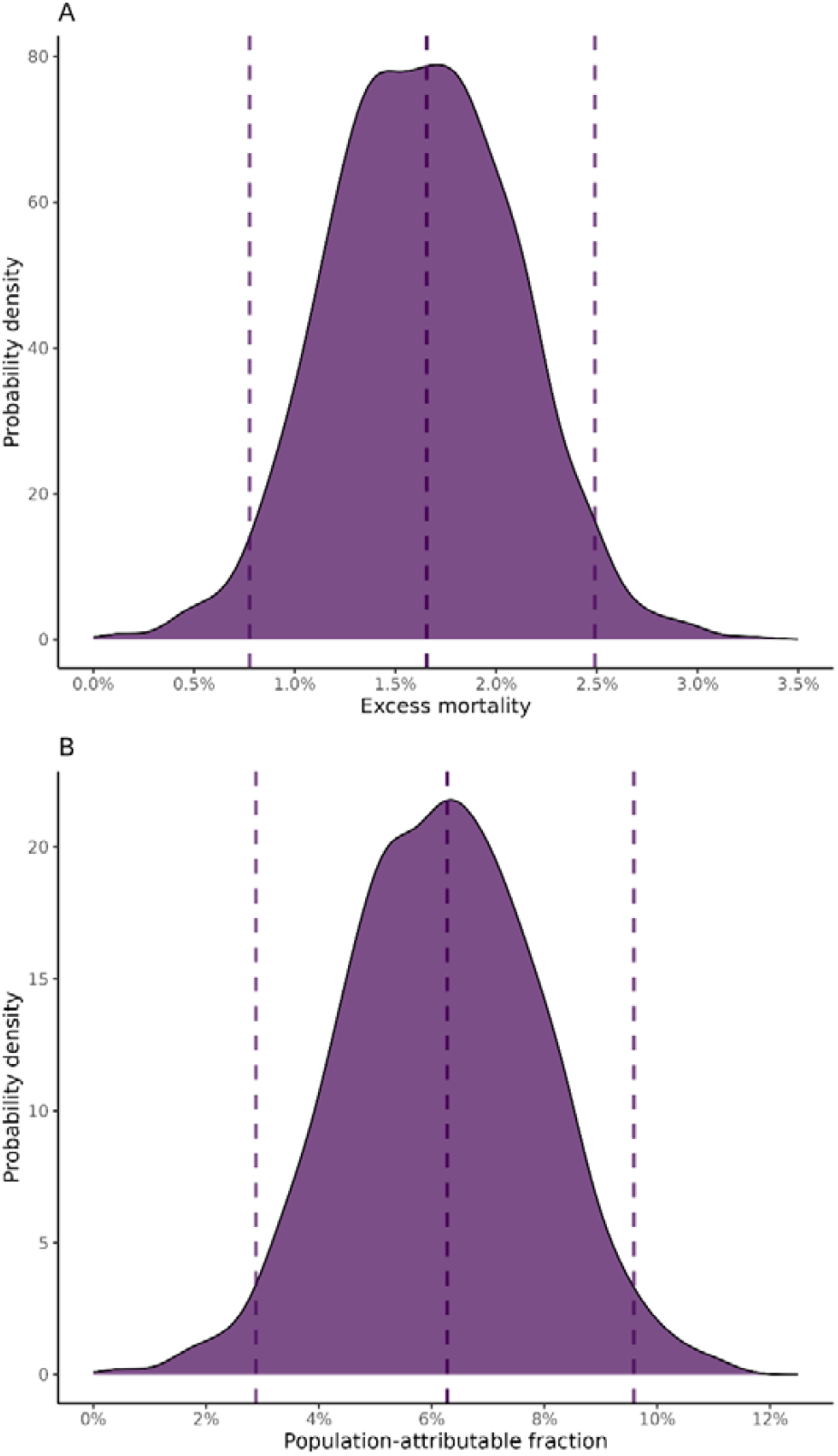
Bootstrapped estimates for cancer patients diagnosed with bacteraemia at Oxford University Hospitals from 2008 to 2018: (A) Density plot of excess mortality estimates with dashed lines indicating the point estimate and confidence interval limits; (B) Density plot of population-attributable fraction estimates, with dashed lines denoting the point estimate and the outer confidence interval limits.

The E-value for excess mortality, converted to a risk ratio, was 1.33 with a lower CI of 1.21, indicating that unmeasured confounders would need to be associated with both the treatment and the outcome by a combined risk ratio of at least 1.33 to explain away the observed association.

## Discussion

In our study investigating the microbial epidemiology of bacteraemias in cancer patients and excess mortality due to AMR at the end of follow-up at Oxford University Hospitals (OxUH) and Oslo University Hospital (OsUH) from 2008-2018, we observed higher resistance proportions in Gram-positive bacteria and *K. pneumoniae*, and more cases of *Acinetobacter* spp. and *P. aeruginosa* at OxUH. Among the 1929 patients at OxUH, 16% had bacteraemia with AMR, while the proportion was 6% of the 1640 patients at OsUH. We estimated an excess mortality of 1.7% and a population-attributable fraction of 6.3% due to AMR at OxUH, while the limited number of AMR events at OsUH (only twelve) precluded a similar estimation. Younger patients at both hospitals showed higher rates of AMR, while solid tumours treated with surgery were most common overall. However, information governance constraints in merging data and the need for patient-level data to control for confounding variables made the calculation of a pooled excess mortality for both locations not feasible.

While VRE were mainly associated with two large outbreaks in Norway and MRSA was still mainly associated with importations, the UK has a higher and more constant background prevalence, indicating some endemicity [18–20]. In Norway and compared to the general population, cancer patients seem to be at a higher risk of contracting VRE due to its healthcare-associated nature, while at a lower risk of MRSA due to its mainly community-associated nature [21]. There is currently a shift in the global hospital epidemiology towards a higher resistance proportions overall and more Gram-negative bacteria with an affinity for acquiring extensive drug-resistance, particularly the non-fermenting organisms *Acinetobacter baumannii* and *P. aeruginosa*, a trend which currently appears to impact the UK and not Norway [22–24].

There have only been a few attempts at estimating the excess mortality attributable to AMR in high income, low-prevalent settings. Moreover, existing studies have not focused on specific high-risk patient groups, such as those with cancer. The relatively low PAF of deaths attributable to AMR at OxUH aligns with the current modest prevalence of AMR in England, suggesting that the problem may not be as advanced as in high-prevalent settings, consistent with estimations from regions facing greater AMR challenges [5]. Although a small difference was observed in the crude mortality at OsUH, we could not estimate the excess mortality due to the small number of events, suggesting the observed difference might have been due to chance. Only about half of studies assessing the impact of AMR on mortality report increased mortality, and mainly from settings with a high prevalence of AMR, yet many fail to find significant impacts. This variation highlights the limited comparability across studies conducted in different contexts. The difficulties may be due to residual confounding or small sample sizes, either due to low prevalence or a focus on specific infectious syndromes or drug-pathogen combinations, challenges that our study also potentially faces [5,25–27]. In low-prevalence AMR settings such as Norway, where AMR cases are sporadic, rarely cause outbreaks, and are often associated with import through tourism or migration [18], individuals infected with resistant microbes may have unique characteristics that are difficult to fully capture in epidemiological analyses. When follow-up was restricted to 7 or 14 days, no differences in the crude mortality were observed between groups at OsUH.

It is important to note that the direct effect of AMR on mortality is likely low – although not zero – as there is overlap between AMR and increased virulence, e.g., due to successful clones or plasmids carrying linked resistance and virulence genes [28]. The impact of AMR on mortality is thus almost exclusively indirect and mediated through the administration of active or inactive antibiotic treatment [29–32]. In both settings, standard empirical regimens differ slightly for patients with a well-managed underlying disease, with amoxicillin/clavulanate and gentamicin commonly used at OxUH and benzylpenicillin and gentamicin at OsUH, while neutropenic patients are often treated with piperacillin/tazobactam. The magnitude of the indirect effect will also depend on the propensity to escalate treatment and the laboratory turnaround time for identifying resistance. If patients expected to have infections by resistant microbes receive empirical treatment to which the microbes are susceptible, we would not expect to observe an increase in mortality. While resistant cases may be overlooked when they are not commonplace, this suggests the benefit of maintaining a low AMR prevalence, allowing healthcare providers to apply extra vigilance and tailored care in suspected AMR cases.

These findings are therefore most likely to be informative for settings with similarly low AMR prevalence and high likelihood of effective empirical treatment.

The quantities we have estimated are intended for a causal interpretation, but such an interpretation is contingent on the assumption that the exposure groups do not differ in unmeasured variables that may confound the relationship between AMR and mortality. The relatively low E-value suggests that even moderate unmeasured confounding could challenge our findings. Several potential confounders remained unmeasured in our study. For example, the type of clinical condition at the time of bacteraemia can influence both the AMR of the causative agent and the mortality. Specifically, the depth and duration of neutropenia can affect the risk of acquiring resistant microbes and directly impact mortality risk. Prior antibiotic exposure is known to select for AMR, and these infections may have a higher risk of leading to adverse outcomes, including death, although a sensitivity analysis adding this covariate did not change our estimate. In addition to the general assumptions for causal inference [13], it is also important to note our assumption that AMR remains constant for 30 days following the onset of bacteraemia. This simplification is practical as it allows for an estimation only involving baseline covariates, but it likely leads to a slight underestimation of true excess mortality due to the potential for either development of AMR or a new infection with AMR during an episode of bacteraemia [33]. We also did not account for whether active or inactive antimicrobial treatment was given, either initially or subsequently. Nevertheless, it is likely that the main driver of mortality was the presence of unexpected AMR not covered by the empiric regimen. Furthermore, our analysis covers only a limited time window providing a snapshot of the total exposure to infectious complications in cancer care, focusing solely on the first bacteraemic episode post-cancer diagnosis as episodes beyond this would be subject to immortal time bias within our study design [34]. The weighted Kaplan-Meier curves demonstrate how excess mortality is inherently time-dependent, suggesting that including multiple episodes or allowing AMR to vary over time would be appropriate, but necessitates more complex time-varying methods.

Our method of estimating the disease burden of AMR differs from some of the most widely cited approaches, which have varying estimation targets and procedures [1–3]. Counterfactual frameworks vary, particularly in the scenarios they contrast, such as all infections being susceptible versus resistant infections replaced by no infections [35,36]. Therefore, our study might lack an upper bound of excess mortality, owing to missing data on individuals without any infection. While direct comparisons with other studies may be challenging [8], our approach offers some advantages as it provides a flexible framework that can be easily adapted to different populations and settings [5].

This study’s strengths include providing insights into AMR in bacteraemias within two distinct healthcare systems, a focus on cancer patients which facilitated a specific theoretical model, and a high data completeness due to routinely collected health data. The research employed a simple and flexible framework for excess mortality estimation, applicable across diverse populations and settings. Nevertheless, there were several limitations in addition to unmeasured confounding discussed above. Our study’s time-fixed counterfactual framework represents a simplification of complex clinical realities and unique characteristics to each data source may hamper comparability. Patients diagnosed with their first cancer pre-2008 might be included with a relapse or metastasis, possibly leading to a higher proportion of patients with later-stage disease at OxUH and early in the study period at both hospitals. On the other hand, patients with cancer may have been treated at OxUH without the ICD-10 code for cancer being set, although this would not impact the results if happening at random. The lack of data on previous antibiotic use in the Norwegian health registry ecosystem poses a challenge for AMR epidemiology studies, although its inclusion in the OxUH cohort did not alter our estimates. While we cannot rule out some misclassification of resistance status or mortality, the use of structured EHR and laboratory data minimises this risk. Any remaining misclassification is likely non-differential and would bias estimates towards the null.

In conclusion, despite identifying only a modest excess mortality due to AMR at OxUH and insufficient data to estimate this at OsUH, there are potential long-term implications of these findings. The microbial distribution was less favourable at OxUH, and the patient case mix also varied in many ways, with younger patients and a higher prevalence of haematological cancers at OxUH. The estimated low excess mortality masks the considerable increase in mortality when the bacteraemia is caused by a resistant phenotype, indicating a challenge that may escalate with a changing microbial epidemiology. Future work should focus on exploring how the effect of AMR varies over time and understanding AMR’s mediation by antibiotic selection.

## Data Availability

The data analysed are not publicly available as they contain personal data. Oxfordshire data are available from the Infections in Oxfordshire Research Database (https://oxfordbrc.nihr.ac.uk/research-themes-overview/antimicrobial-resistance-and-modernising-microbiology/infections-in-oxfordshire-research-database-iord/), subject to an application and research proposal meeting on the ethical and governance requirements of the Database. Data from the Norwegian health registries can be accessed via https://helsedata.no/en/ following an application process, with guides available on the site detailing the requirements and steps for access.

## Acknowledgements

This work uses data provided by patients and collected by the UK’s National Health Service as part of their care and support. We thank all the people of Oxfordshire who contribute to the Infections in Oxfordshire Research Database. Research Database Team: L Butcher, H Boseley, C Crichton, DW Crook, D Eyre, O Freeman, J Gearing (community), R Harrington, K Jeffery, M Landray, A Pal, TEA Peto, TP Quan, J Robinson (community), J Sellors, B Shine, AS Walker, D Waller. Patient and Public Panel: G Blower, C Mancey, P McLoughlin, B Nichols. We would also like to thank the SE Amdal and M Lien for their data capture from the microbiological laboratory information systems in Oslo, and the research group at the Big Data Institute for all the discussions. Data from the Cancer Registry of Norway and the Norwegian Patient Registry has been used in this research, but interpretation and reporting are our sole responsibility, and no endorsement by these registries is intended nor should be inferred.

## Funding

This study was funded by a grant from the South-Eastern Norway Regional Health Authority trust (grant number 2021013), and by the National Institute for Health Research Health Protection Research Unit (NIHR HPRU) in Healthcare Associated Infections and Antimicrobial Resistance at Oxford University in partnership with the UK Health Security Agency (NIHR200915), and the NIHR Biomedical Research Centre, Oxford. DWE is a Big Data Institute Robertson Fellow. The views expressed are those of the authors and not necessarily those of the NHS, the NIHR, the Department of Health or the UK Health Security Agency. The funders had no role in study design, data collection and analysis, decision to publish, or preparation of the manuscript.

## Conflicts of interest

Nothing to declare.

## Tables and figures

**Figure S1.**
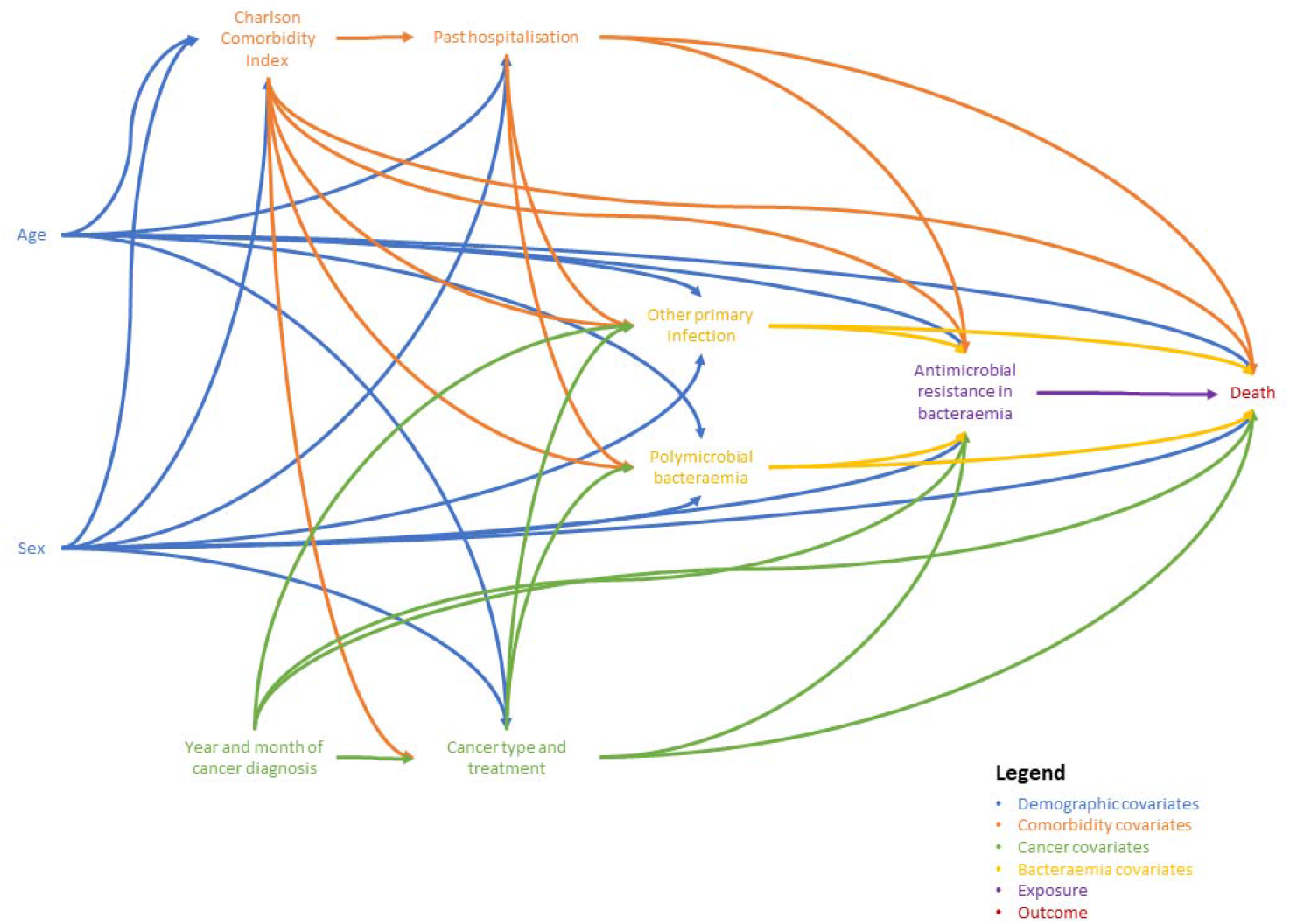
Directed acyclic graph.

## Supplementary material

Supplementary material S2 – Detailed covariate description

- Sex: Recorded as either female or male.
- Age: Treated as a continuous variable, derived from the date of birth.
- Charlson Comorbidity Index (CCI): Calculated from ICD-10 diagnosis codes within one year prior to the bacteraemia episode.
- Past hospitalisation: The cumulative sum of all hospital stay days within one year prior to the bacteraemia episode, reflecting previous exposure to secondary healthcare, but not to long-term care facilities or primary healthcare. “Patient hotels” are counted as inpatient stays in Norway, and the logistical challenges posed by Norway’s geography combined with OsUH’s national functions might contribute to differences in hospitalisation days.
- Other infectious focus: Identified by the presence of infectious syndromes in other sites than blood (thus excluding septicaemia). As the specimen was not always known at OsUH if not blood, we relied only on ICD-10 codes at both hospitals, basing our classifications of the focus on the Clinical Classifications Software Refined [37].
- Year and month of cancer diagnosis: Used to adjust for time-related changes in cancer treatment and supportive care.
- Cancer type and treatment: Categorised based on malignancy and treatment within a year before follow-up, including solid cancers with or without surgery and haematological cancers with or without stem cell transplantation. To classify surgeries, a concept-based approach was adopted in which a list of common terms for surgeries in both English and Norwegian were used to search in the procedure code descriptions. To capture exposures related to transplantation, a 90-day lookahead was also adopted, as transplantations were sometimes planned with (myeloablative and/or immunosuppressive) conditioning started, without the procedure having been completed. This coding was not perfect, however, due to immortal time bias.
- Polymicrobial bacteraemia: A binary variable defined by the identification of different causative agents in the same or adjacent days’ blood cultures, with coagulase-negative staphylococci generally considered contamination. Please note that differences in case mix, including OsUH’s role as Norway’s specialist cancer centre, may account for variations in patient age and polymicrobial bacteraemias due to diverse underlying immunosuppression, among other undetermined factors.

All information from OxUH comes from the hospital’s electronic health record system, coded and stored in the Infections in Oxfordshire Research Database. Sex, age, date of cancer diagnosis and cancer type at OsUH was collected from the Cancer Registry of Norway, past hospitalisation, ICD-10 codes, and procedure codes were collected from the Norwegian Patient Registry, and all microbiological data were collected from the raw data of the microbiological laboratory information systems.

### Supplementary material S3 – Detailed statistical methods

We consider the excess mortality due to AMR to be the difference between the actual observed mortality and the expected mortality at the end of follow-up had all bacteraemia been caused by susceptible bacteria. As the latter is a counterfactual number that is not observed directly and needs to be estimated, excess mortality is most precisely defined using counterfactual notation. The resulting estimand may be described as

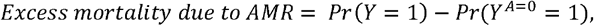

where *Y* is the outcome variable indicating mortality, with *Y* = 1 representing death and *Y* = 0 indicating survival at the end of follow-up. A represents the exposure, with *A* = 1 indicating a bacteraemia with a resistant phenotype and *A* = 0 indicating a susceptible phenotype. Thus, *Pr*(*Y* = 1) represents the observed mortality in the population, and *Pr(Y*^*A*=0^ = 1) represents the expected mortality if all participants had a bacteraemia caused by a susceptible phenotype. We will also estimate the population-attributable fraction (PAF), which we define as

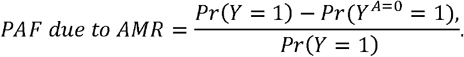

PAF is inconsistently defined in epidemiological literature [6], our definition was based on our definition of excess mortality and consisted of contrasting the observed and the expected mortalities at the end of follow-up.

To estimate the expected mortality in the absence of AMR, i.e. *Pr(Y*^*A=0*^ = 1), we weighted the bacteraemias without AMR by the inverse probability of not having AMR, conditioning on the pre-identified confounders [13,14]. Under the mentioned identification assumptions, this weighted subpopulation is representative of a full population without AMR. We calculated stabilised, untruncated inverse probability weights using logistic regression, with the confounders as independent variables and the exposure group as the dependent variable. Covariates were included as outlined in the methods chapter. Continuous variables were modelled linearly, as more complex functional forms offered only marginal improvements in model fit based on Akaike’s information criterion. The expected cumulative incidence function of mortality was then subtracted from the observed cumulative incidence function to obtain the excess mortality estimate. We then used nonparametric bootstrapping with 1000 bootstrap samples to calculate percentile-based 95% confidence intervals for both excess mortality and PAF [13]. This approach required a certain sample size as insufficient events in both groups to ensure model stability and convergence may render bootstrapping and estimation procedures unfeasible.

We then estimated the unweighted Kaplan-Meier survival curves by exposure groups, and the unweighted survival curve for the overall observed mortality with the weighted survival curve for the counterfactual scenario with confidence intervals bootstrapped at each time point. This would correspond to defining excess mortality as the difference between survival functions *S*(t) - *S*^*A=*0^ (t) effectively quantifying it day-by-day in our weighted analysis. Density plots then illustrated the excess mortality and PAF at the end of follow-up with the bootstrapped confidence intervals. Excess mortality was also estimated for each drug-pathogen combination separately and a sensitivity analysis was performed for the OxUH data adding a linear term for cumulative days on antibiotics in the previous 365 days. Lastly, the E-value was calculated to evaluate the minimum strength of association required for an unmeasured confounder to fully explain away the estimated excess mortality due to AMR, under the assumption that measured covariates were correctly controlled for; this was done by first converting the risk difference to a risk ratio by dividing the counterfactual and excess mortality by the counterfactual mortality [15].

A causal interpretation of the estimates in our study relies on three central identifiability conditions being met: consistency, exchangeability, and positivity [13].

1. **Consistency:** This condition asserts that the observed outcome for an individual under their actual exposure is the same as the estimated outcome if that patient happened to be ascribed the same counterfactual exposure. For example, if a patient had a susceptible phenotype, their estimated outcome in the counterfactual scenario where all bacteraemias were caused by susceptible phenotypes should be identical to their observed outcome. This means the treatment and outcome are well-defined and consistently applied.
2. **Exchangeability:** Also known as no unmeasured confounding, this condition posits that the groups being compared are similar in all relevant respects except for the exposure of interest. It necessitates a correct adjustment set where all known confounding variables are accounted for without introducing bias by conditioning on certain covariates known as colliders. This mimics the randomisation in controlled trials. In our context, we assume that by controlling for all identified confounders (as informed by our DAG), the exposure groups (resistant vs. susceptible phenotypes) are exchangeable. However, unmeasured confounding remains a concern, as it is challenging to be certain that our DAG is completely accurate. Sensitivity analyses and the calculation of the E-value, which assesses the robustness of our findings to potential unmeasured confounders, address this limitation such that it can be assessed whether the assumption of exchangeability is reasonable.
3. **Positivity:** This condition assumes that every individual has a non-zero probability of receiving the exposure within each level of every included covariate. This is crucial to avoid biases in effect estimation due to a limited data range. In our study, positivity was ensured by the selection of the patient population and the coding of covariates, such as combining cancer types and treatments. We ensured there were both resistant and susceptible phenotypes across all levels of our covariates, reducing the risk of violating this assumption.

**Table S3.** The unweighted and weighted mean values of the continuous and dichotomous covariates included in the propensity score model used to weight the population in the counterfactual scenario of absence of antimicrobial resistance in all bacteraemias among cancer patients at Oxford University Hospitals, 2008-2018.

|  | **Mean values** |  |
| --- | --- | --- |
|  | **AMR**<br>**absent** | **AMR**<br>**present** |
| **Unweighted** |  |  |
| Age | 66.23 | 62.95 |
| Sex | 0.61 | 0.69 |
| Other primary | 0.50 | 0.42 |
| Polymicrobial | 0.04 | 0.04 |
| Previous hospitalisation | 27.04 | 41.47 |
| **Weighted** |  |  |
| Age | 65.74 | 65.26 |
| Sex | 0.63 | 0.65 |
| Other primary | 0.49 | 0.50 |
| Polymicrobial | 0.04 | 0.04 |
| Previous hospitalisation | 29.60 | 30.58 |

**Table S4.**
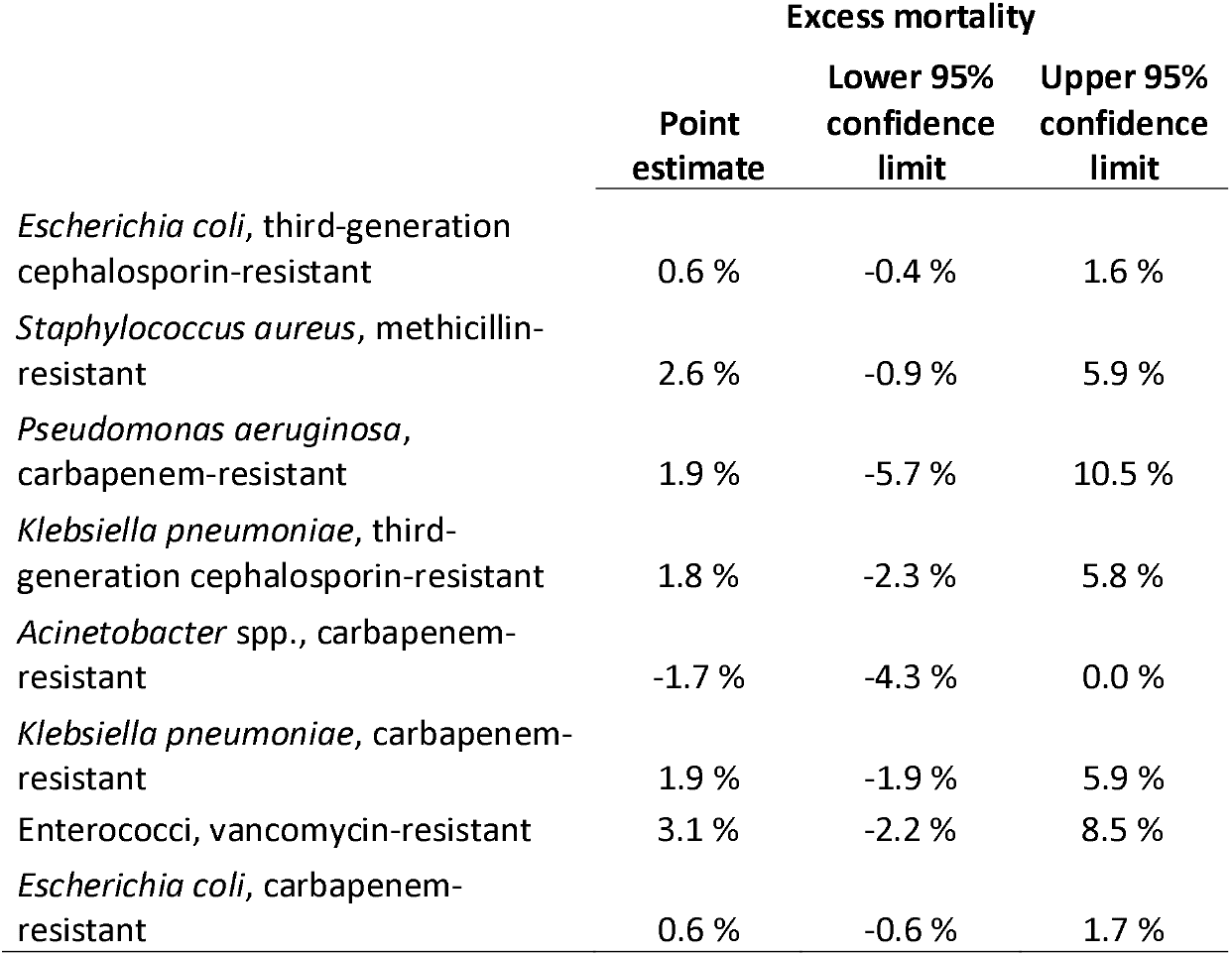
Excess mortality estimates and bootstrapped 95% confidence intervals of head-to-head comparisons of the key drug-pathogen combinations isolated from the blood of cancer patients diagnosed in 2008-2018 at Oxford University Hospitals.

|  | Excess mortality |  |  |
| --- | --- | --- | --- |
|  | Point estimate | Lower 95% confidence limit | Upper 95% confidence limit |
| *Escherichia coli*, third-generation cephalosporin-resistant | 0.6 % | -0.4 % | 1.6 % |
| *Staphylococcus aureus*, methicillin-resistant | 2.6 % | -0.9 % | 5.9 % |
| *Pseudomonas aeruginosa*, carbapenem-resistant | 1.9 % | -5.7 % | 10.5 % |
| *Klebsiella pneumoniae*, third-generation cephalosporin-resistant | 1.8 % | -2.3 % | 5.8 % |
| *Acinetobacter* spp., carbapenem-resistant | -1.7 % | -4.3 % | 0.0 % |
| *Klebsiella pneumoniae*, carbapenem-resistant | 1.9 % | -1.9 % | 5.9 % |
| Enterococci, vancomycin-resistant | 3.1 % | -2.2 % | 8.5 % |
| *Escherichia coli*, carbapenem-resistant | 0.6 % | -0.6 % | 1.7 % |

**Table S5.**
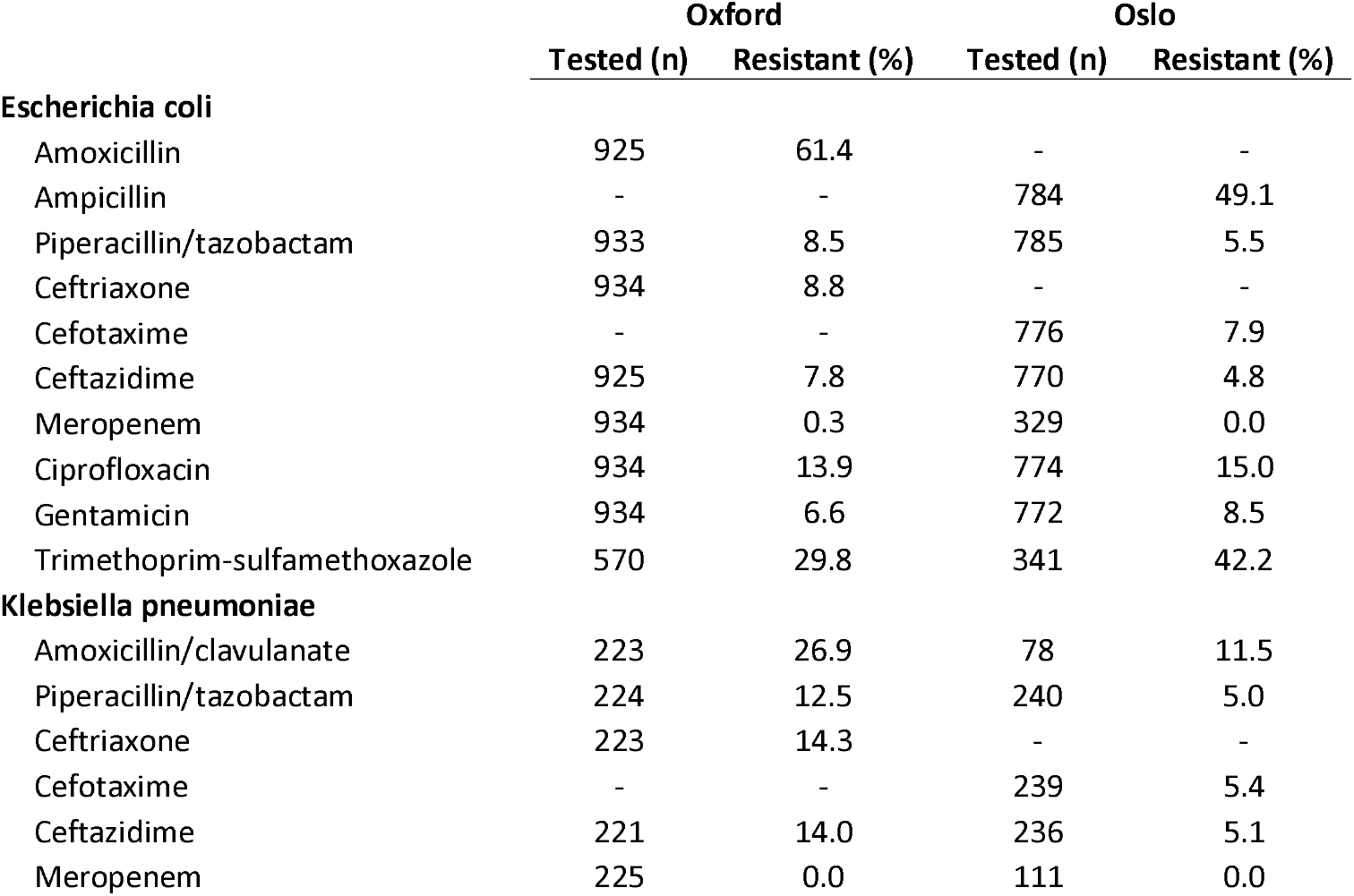
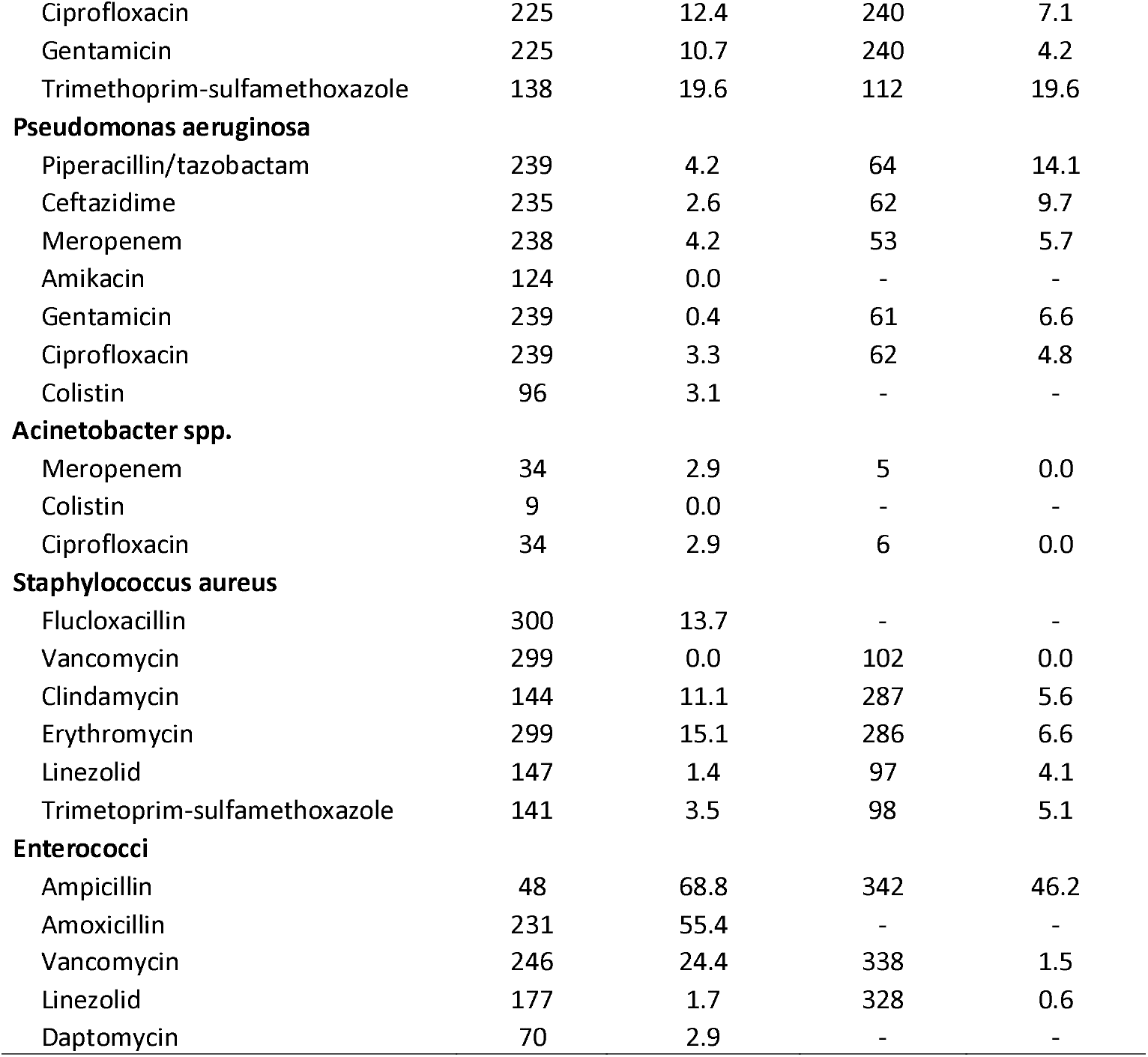
Cumulative antibiogram of the included microbes isolated from the blood of cancer patients diagnosed in 2008-2018 at Oxford University Hospitals and Oslo University Hospital.

|  | Oxford |  | Oslo |  |
| --- | --- | --- | --- | --- |
|  | Tested (n) | Resistant (%) | Tested (n) | Resistant (%) |
| **Escherichia coli** |  |  |  |  |
| Amoxicillin | 925 | 61.4 | - | - |
| Ampicillin | - | - | 784 | 49.1 |
| Piperacillin/tazobactam | 933 | 8.5 | 785 | 5.5 |
| Ceftriaxone | 934 | 8.8 | - | - |
| Cefotaxime | - | - | 776 | 7.9 |
| Ceftazidime | 925 | 7.8 | 770 | 4.8 |
| Meropenem | 934 | 0.3 | 329 | 0.0 |
| Ciprofloxacin | 934 | 13.9 | 774 | 15.0 |
| Gentamicin | 934 | 6.6 | 772 | 8.5 |
| Trimethoprim-sulfamethoxazole | 570 | 29.8 | 341 | 42.2 |
| **Klebsiella pneumoniae** |  |  |  |  |
| Amoxicillin/clavulanate | 223 | 26.9 | 78 | 11.5 |
| Piperacillin/tazobactam | 224 | 12.5 | 240 | 5.0 |
| Ceftriaxone | 223 | 14.3 | - | - |
| Cefotaxime | - | - | 239 | 5.4 |
| Ceftazidime | 221 | 14.0 | 236 | 5.1 |
| Meropenem | 225 | 0.0 | 111 | 0.0 |
| Ciprofloxacin | 225 | 12.4 | 240 | 7.1 |
| Gentamicin | 225 | 10.7 | 240 | 4.2 |
| Trimethoprim-sulfamethoxazole | 138 | 19.6 | 112 | 19.6 |
| **Pseudomonas aeruginosa** |  |  |  |  |
| Piperacillin/tazobactam | 239 | 4.2 | 64 | 14.1 |
| Ceftazidime | 235 | 2.6 | 62 | 9.7 |
| Meropenem | 238 | 4.2 | 53 | 5.7 |
| Amikacin | 124 | 0.0 | - | - |
| Gentamicin | 239 | 0.4 | 61 | 6.6 |
| Ciprofloxacin | 239 | 3.3 | 62 | 4.8 |
| Colistin | 96 | 3.1 | - | - |
| **Acinetobacter spp.** |  |  |  |  |
| Meropenem | 34 | 2.9 | 5 | 0.0 |
| Colistin | 9 | 0.0 | - | - |
| Ciprofloxacin | 34 | 2.9 | 6 | 0.0 |
| **Staphylococcus aureus** |  |  |  |  |
| Flucloxacillin | 300 | 13.7 | - | - |
| Vancomycin | 299 | 0.0 | 102 | 0.0 |
| Clindamycin | 144 | 11.1 | 287 | 5.6 |
| Erythromycin | 299 | 15.1 | 286 | 6.6 |
| Linezolid | 147 | 1.4 | 97 | 4.1 |
| Trimetoprim-sulfamethoxazole | 141 | 3.5 | 98 | 5.1 |
| **Enterococci** |  |  |  |  |
| Ampicillin | 48 | 68.8 | 342 | 46.2 |
| Amoxicillin | 231 | 55.4 | - | - |
| Vancomycin | 246 | 24.4 | 338 | 1.5 |
| Linezolid | 177 | 1.7 | 328 | 0.6 |
| Daptomycin | 70 | 2.9 | - | - |

## Notes

### Competing Interest Statement

The authors have declared no competing interest.

### Author Declarations

Data from OxUH was obtained from the Infections in Oxfordshire Research Database (IORD) with approvals from the National Research Ethics Service South Central-Oxford C Research Ethics Committee (19/SC/0403), Health Research Authority and Confidentiality Advisory Group (19/CAG/0144) as a deidentified database without individual consent. The Regional Ethics Committee of South East Norway (240258) and the Data Protection Officer at OsUH (21/06874) gave ethical approval for the work involving Norwegian registry data.

### Summary of Updates

Revised some of the text in the manuscript and included a cumulative antibiogram.

